# Prevalence of Non-alcoholic Fatty Liver Disease (NAFLD) among Children and Adolescents (<18 years) in India: A Systematic Review and Meta-analysis

**DOI:** 10.64898/2026.08.12.26360221

**Authors:** MD. Abu Bashar, Prabhat, Imran Ahmed Khan, Nazia Begam

**Author notes:** Joint first authors (MD. Abu Bashar, Prabhat).

## Abstract

**Background:** Non-alcoholic fatty liver disease (NAFLD) has become a common metabolic disorder in paediatric age groups and is a major contributor to the burden of health economics. However, there is a lack of comprehensive data on the prevalence of this condition among children.

**Methods:** English language literature published from inception until April 2025 was searched from the electronic databases, i.e., PubMed/Medline, Scopus, Embase, and CINAHL. Original data published in any form that reported NAFLD prevalence among children and/or adolescent population from India were included. The subgroup analysis of prevalence was done based on the risk category, i.e., average-risk group, and high-risk group (obesity or overweight, metabolic syndrome, etc.). The prevalence estimates were pooled using the random-effects model.

**Results:** A total of 11 studies (six in high-risk populations and 5 in the average-risk general population) comprising data from 3512 individuals were found eligible and were included. The overall pooled estimate of NAFLD prevalence among the children and adolescents was 35.4% (95% CI: 19.7%–52.9%) with very high heterogeneity(I^2^=99.0%). The pooled prevalence of NAFLD among **average/low risk** children and adolescents was 10.7% (95%CI: 5.2% to 20.5%) with high heterogeneity across the studies (I^2^= 96.6%, p=0.001) whereas the pooled prevalence of NAFLD in high risk overweight/obese children and adolescents was found to be 59.7% (95% CI:55.2% to 64.1%) with moderate heterogeneity across the studies (I^2^=50.90%; H^2^=2.04; Q (5) = 10.03; p=0.07)

**Conclusion:** This systematic review demonstrates that non-alcoholic fatty liver disease (NAFLD) poses a growing health concern among Indian children and adolescents, as 1 out of 3 children/adolescents are suffering from it, with a disproportionately high burden observed in those who are overweight or obese.

## Introduction

Non-alcoholic fatty liver disease (NAFLD) is characterized by the accumulation of excessive fat in the liver. In the later stages of the disease, inflammation occurs, and subsequently hepatocyte injury and fibrosis, leading to non-alcoholic steatohepatitis (NASH) (1). NAFLD is diagnosed by imaging studies and histopathology after excluding the known causes of hepatic fat accumulation, such as chronic alcoholism, use of steatogenic medication, or hereditary disorders (2). The global prevalence of NAFLD is reported to be 25%, and that of NASH is 1.5%–6.5% (3). NAFLD was previously thought to be a disease of adults only, but subsequently, a significant rise in paediatric NAFLD cases was observed, associated with a rising prevalence of childhood obesity and metabolic syndrome (4,5). Rapid urbanization, increasing sedentary behaviour, dietary habits, and genetic predisposition have contributed to an alarming rise in childhood obesity, which is a precursor to NAFLD (6).

Studies from various parts of the world have reported a varying range for the prevalence of NAFLD from 7.4% to 23.8%% % in the healthy population of children and adolescents (7, 8). The global NAFLD prevalence in adolescents increased from 3.73% to 4.71% from 1990 to 2019 (a relative increase of 26.27%) (9). As per a systematic review, the prevalence of pediatric NAFLD is 7.6 % in the general population and 34.2% in the clinically obese population (10).

Although NAFLD is usually considered to be a benign disease, its prevalence increases with age (11). Further, the prevalence of prediabetes, diabetes, and metabolic syndrome is increasing in India, both in urban and rural populations, which are associated with NAFLD (12, 13). The high prevalence of NAFLD, along with diabetes, obesity, and metabolic syndrome in the Indian population, has significantly increased the burden on healthcare resources (14). Moreover, to address this public health problem, a detailed analysis of the recent prevalence and a plan with proper future strategies must be created and implemented. There are multiple studies on NAFLD from India, but most have limitations such as small sample sizes, regional variation, bias, improper definition of populations, etc. Moreover, the studies also lack proper segregation into low/high-risk groups (15–17).

There is a paucity of data regarding the prevalence of NAFLD among children and adolescents from India. The estimated burden varies among different studies due to the regional differences in disease prevalence, variability in study methodologies, and diagnostic criteria utilized. A systematic synthesis of available data is essential to better understand the magnitude of NAFLD among children to identify at-risk populations and to tailor public health strategies for early detection and prevention. This systematic review and meta-analysis aimed to estimate the pooled prevalence of NAFLD among children and adolescents (aged<18 years) in India and to stratify it by risk categories.

### Primary objective

**1.** To estimate the pooled prevalence of NAFLD among Indian children and adolescents (< 18 years)

### Secondary objective

**1.** To compare the prevalence of NAFLD among children who are at low-risk/average-risk with those at high-risk for developing NAFLD.

## Material & Methods

### Design

We followed the Preferred Reporting Items for Systematic Reviews and Meta-Analyses (PRISMA) checklist for conducting the study (18).

### Search Strategy

We searched electronic databases including PubMed/Medline, Embase, Scopus, CINAHL, and Google Scholar. The search strategy (Supplementary file 1) included the various terms used for fatty liver disease, the name of states, and major cities of the country. Cross-references from the published articles were manually searched to retrieve the additional literature.

### Inclusion and Exclusion Criteria

We included English language literature published as full text till April 2024. The studies were included if they reported original data on the prevalence of fatty liver disease in the Indian children/ adolescent population in any form, such as original articles, letters to the editor, brief communications, or short reports. We excluded abstracts, review articles, and non-English language literature. The studies that reported NAFLD prevalence in India based on ultrasound as the imaging modality were selected for data extraction and analysis. In studies reporting the prevalence of NAFLD based on multiple modalities, including ultrasound, we included the reported prevalence based on the latter modality.

### Study Participants

We included children and adolescents aged up to 18 years. We also included studies reporting NAFLD prevalence in high-risk populations such as those with overweight/obesity and metabolic syndrome.

### Selection of Studies

The literature search was performed by PP. Two independent reviewers (MAB and IAK) screened the titles and abstracts of all studies identified. Full-text articles were obtained for the relevant studies satisfying the inclusion criteria. The data were extracted independently by authors PP and MAB. Extracted data were cross-checked by an independent reviewer, IAK. The data extraction was supervised by IAK, and any disagreement between the authors was resolved by consensus.

### Data Extraction

The following data were extracted from the studies: author name, year of publication, study design, sample size, age groups (< 5, 5-10 years, 10-18 years) of the participants, study setting, number of study centres, characteristics of the study population, risk category of the participants, residence, and diagnostic criteria used for the diagnosis of NAFLD. The study population was classified as average-risk (general population, participants of the control arm, unselected participants) or high-risk (obesity or overweight, metabolic syndrome).

### Quality Assessment of the Studies

The quality of the included studies was assessed with the use of the Modified Joanna Briggs Institute (JBI) Checklist (19) and New Castle Ottawa Scale for cross sectional studies (20). The JBI checklist includes a set of eight questions on different methodological quality parameters of a prevalence study. The response to each of the questions was marked as either “Yes” or “No/unclear”. The overall quality of each of the studies was assessed as poor quality, average quality, and high quality.

### Statistical Analysis

The NAFLD prevalence data from individual studies were summarized as proportions with 95% confidence intervals (CIs). The heterogeneity between studies was assessed with I^2^ statistics. The presence of substantial heterogeneity was adjudged using the I^2^ statistic (I^2^ ≥ 50%). The prevalence estimates from individual studies were pooled with a random-effects model because of marked heterogeneity among studies. Publication bias was assessed using Egger’s test with funnel plots. The data were analyzed with STATA software, version 16 (StatCorp LLC, College Station, TX, USA). Subgroup analyses were performed for age group, gender, and risk category.

### Ethical consideration

An ethical review does not apply to this study because it uses data available in the published literature.

### Protocol registration

The SRMA protocol as registered with PROPSERO with registration no. CRD420251026426, available at https://www.crd.york.ac.uk/PROSPERO/view/CRD420251026426

## Results

### Screening Strategy

The search retrieved 802 publications, of which 11 are included in this systematic review, corresponding to 11 independent study populations (Figure 1).

**Fig. 1:**
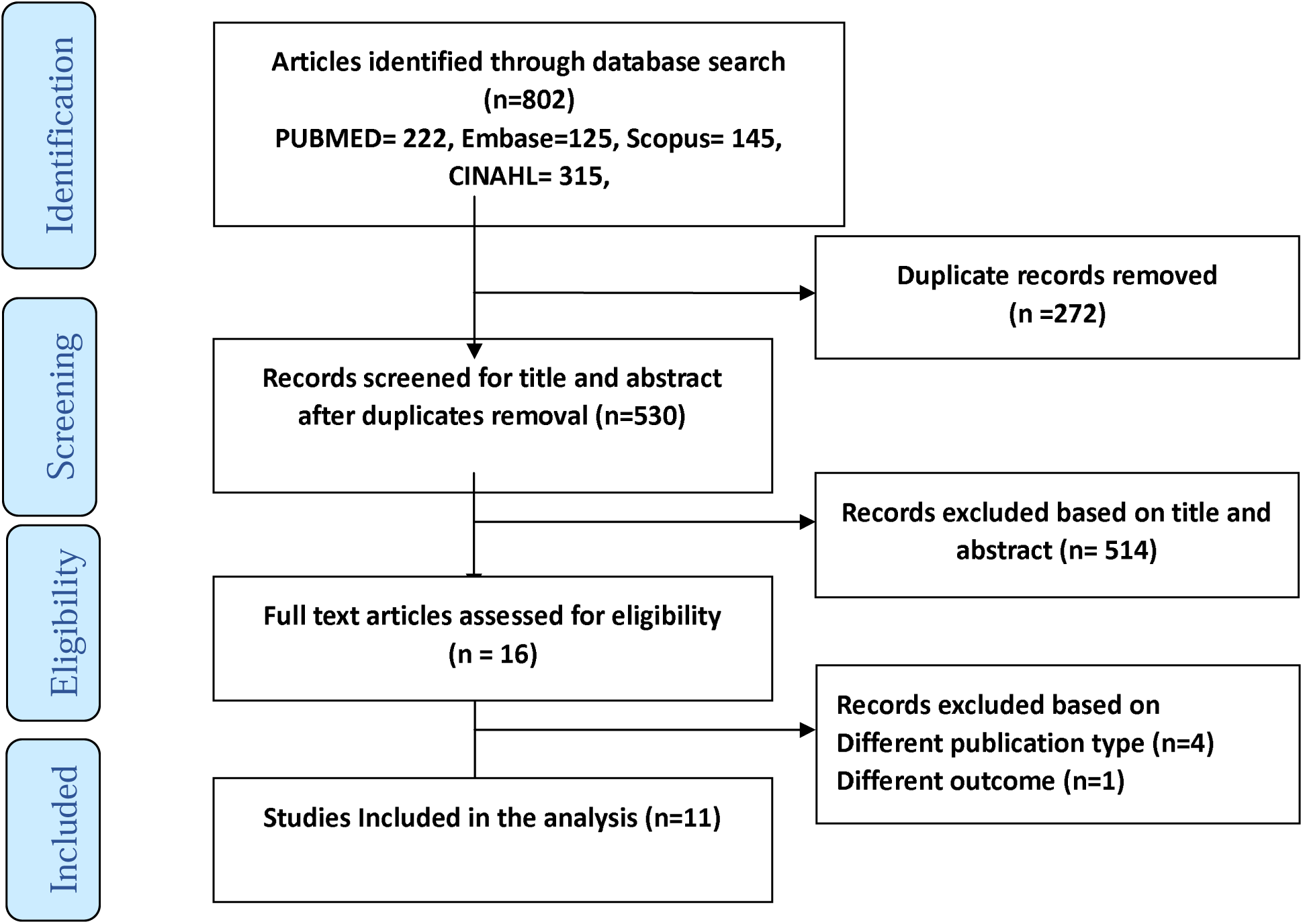
PRISMA flow diagram showing the study selection process.

### Characteristics of included studies

The characteristics of the included studies are summarized in Table 1 (2,20–27). The included studies summarized the data from 3512 participants grouped into 11 datasets. The sample sizes ranged from 100 to 1114, with a median of 160. The oldest study was published in 2012, and the latest in 2023. The main aim of the majority of studies was to screen for NAFLD among high-risk overweight and obese children and to find its predictors. Overall, 5 studies were conducted in average-risk general populations and 6 studies in high-risk populations of overweight/obese children/adolescents. Although the prevalence estimates extracted for this systematic review were reported by studies of various designs (e.g. cross cross-sectional and prospective studies), prevalence estimates themselves are, by nature, cross-sectional.

**Table 1.**
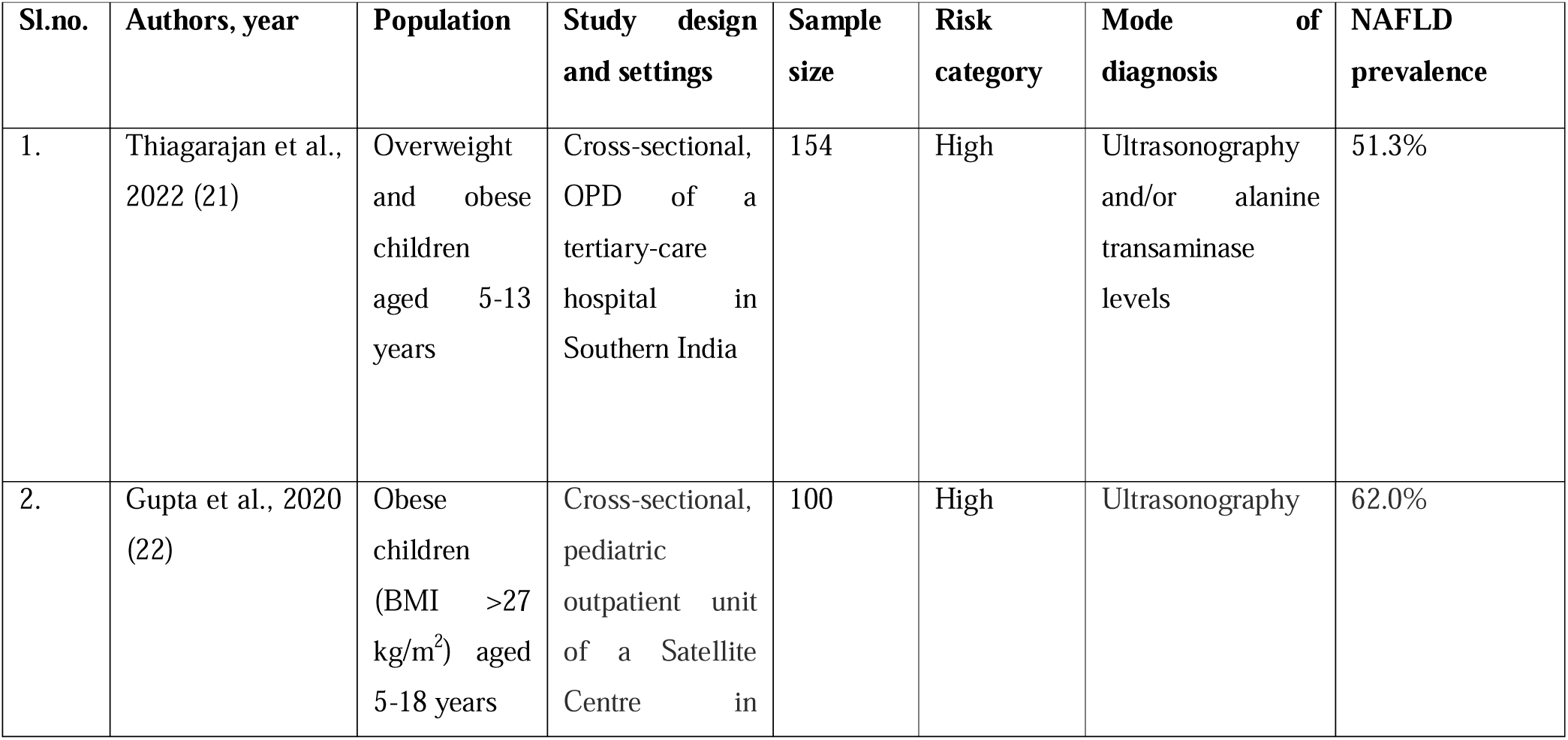
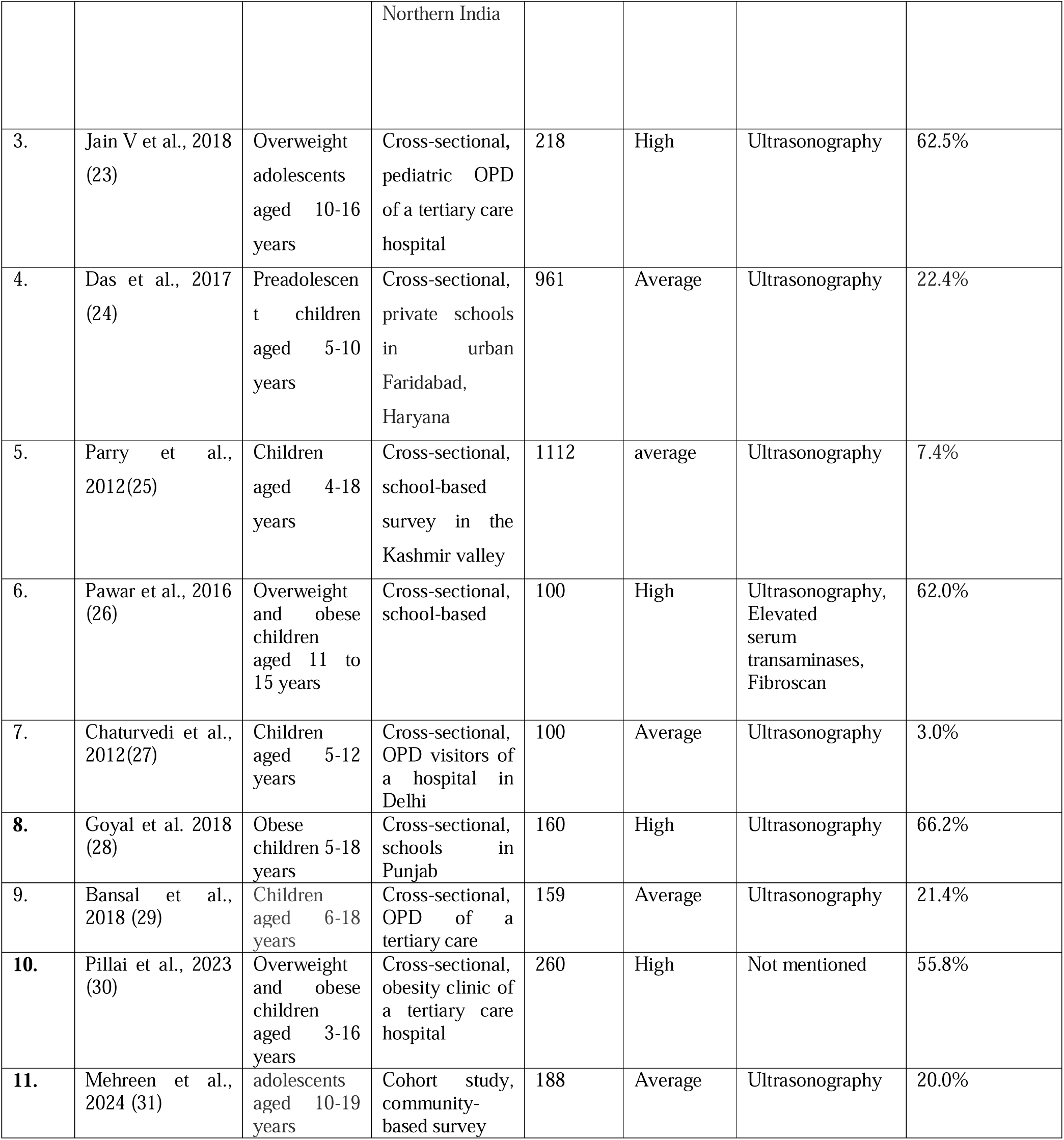
Characteristics of the studies included in the Review.

Eight out of the 11 included studies mentioned Ultrasonography (USG) only as the modality for diagnosing NAFLD, two studies used more than one method, i.e., USG along with serum alanine transaminase (ALT) levels and fibro scan, whereas only one study didn’t mention the modality for diagnosing NAFLD.

### Overall pooled prevalence of NAFLD among children/adolescents

The meta-analysis included data from 3512 children/adolescents aged from 4 to 19 years, of whom 938 had NAFLD (Table 1). All the 11 included studies were single-centre studies and were conducted in 9 states/UTs (Delhi 2, Punjab-2, and 1 each from Himachal Pradesh, Tamil Nadu, Haryana, Maharashtra, Kerala, Puducherry, & Jammu & Kashmir).

The overall pooled estimate of NAFLD prevalence among the children and adolescents was 35.0% (95% CI: 20.0%–52.9%) (Fig. 3) with considerable heterogeneity(I^2^ 99.0%, p=0.00) between the studies.

**Fig. 3.**
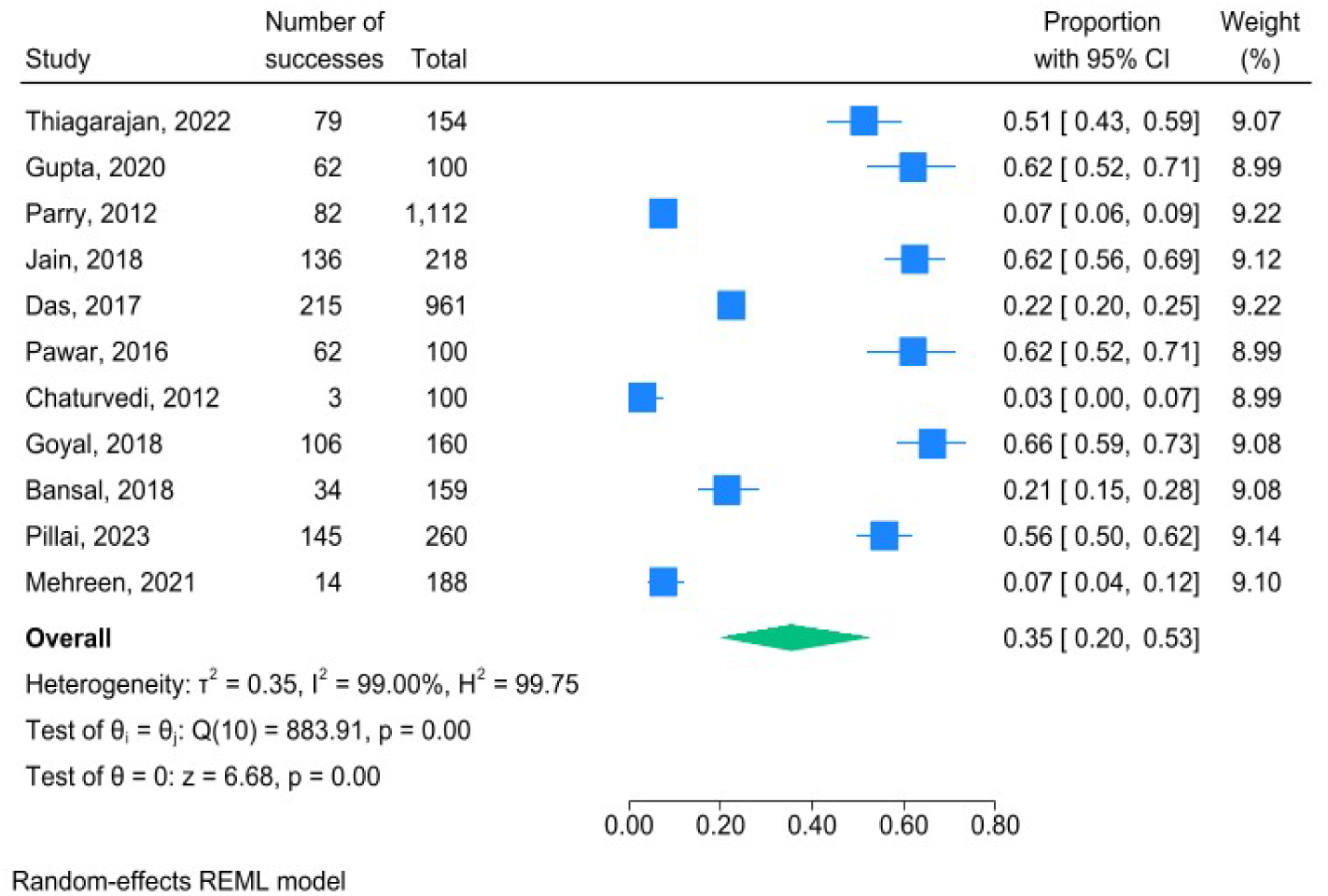
Pooled estimates of NAFLD prevalence by the random-effects model among children and adolescents(n-=11)

### Pooled prevalence of NAFLD among average-risk children/adolescents

Five out of 11 studies were conducted on children/adolescents with an average risk of NAFLD, i.e., children/adolescents from the general population. The overall pooled prevalence of NAFLD in average risk children/adolescents’ studies was 10.7% (95% CI: 5.2% to 20.5%, I^2^= 96.6%, p=0.00 Fig. 4). The I^2^ value of 96.6% and prediction interval ranged from % to %, reflecting the significant between-study heterogeneity.

**Fig. 4.**
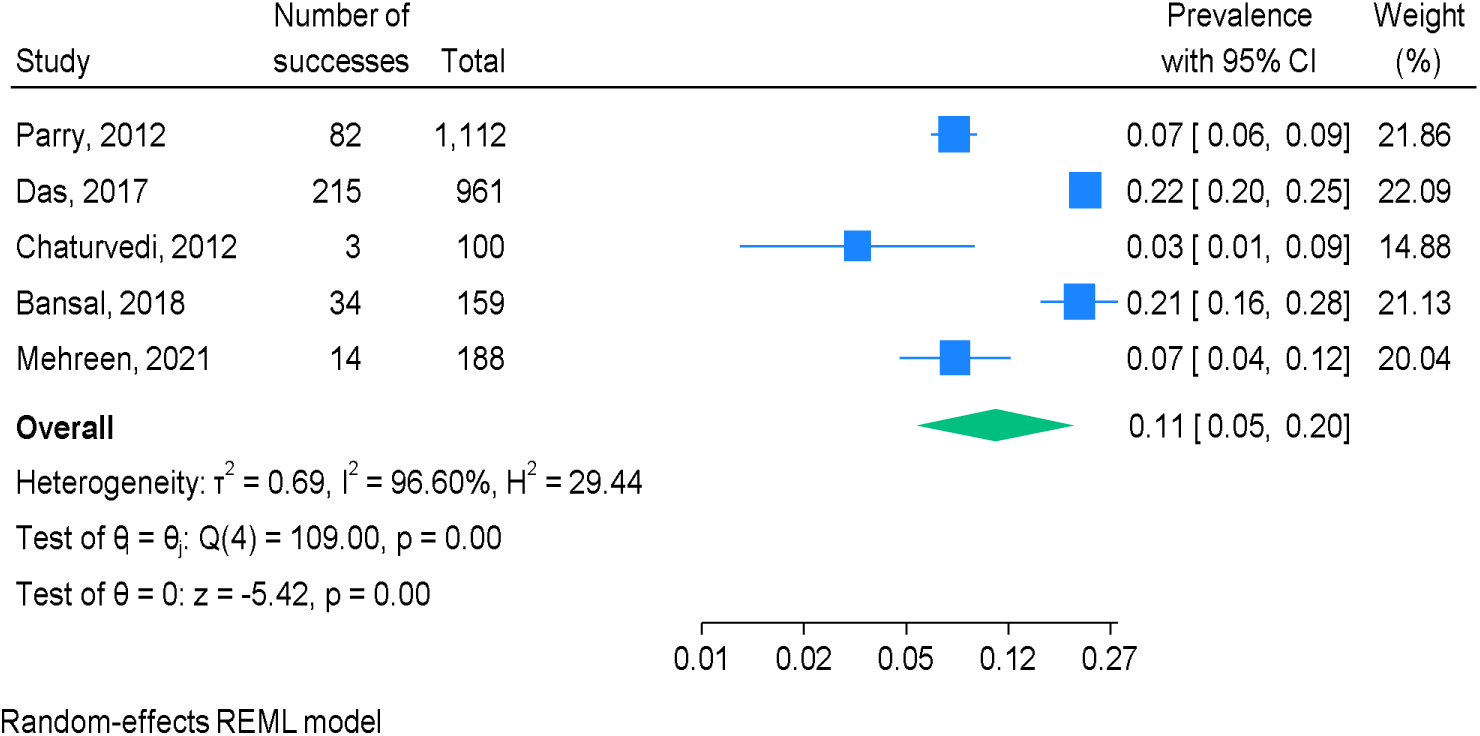
Pooled estimate of NAFLD prevalence among children/adolescents with average-risk(n=5)

### Pooled prevalence of NAFLD among High-risk children/adolescents

Six of the eleven studies reported NAFLD prevalence rates in children/adolescents with a high risk of NAFLD, i.e., overweight/obese population. The pooled prevalence of NAFLD in this subgroup was found to be 59.7% (95% CI:55.2% to 64.1%) with moderate heterogeneity across the studies (I^2^=50.90%; H^2^=2.04; Q (5) = 10.03; p=0.07) (Fig.5).

**Fig. 5.**
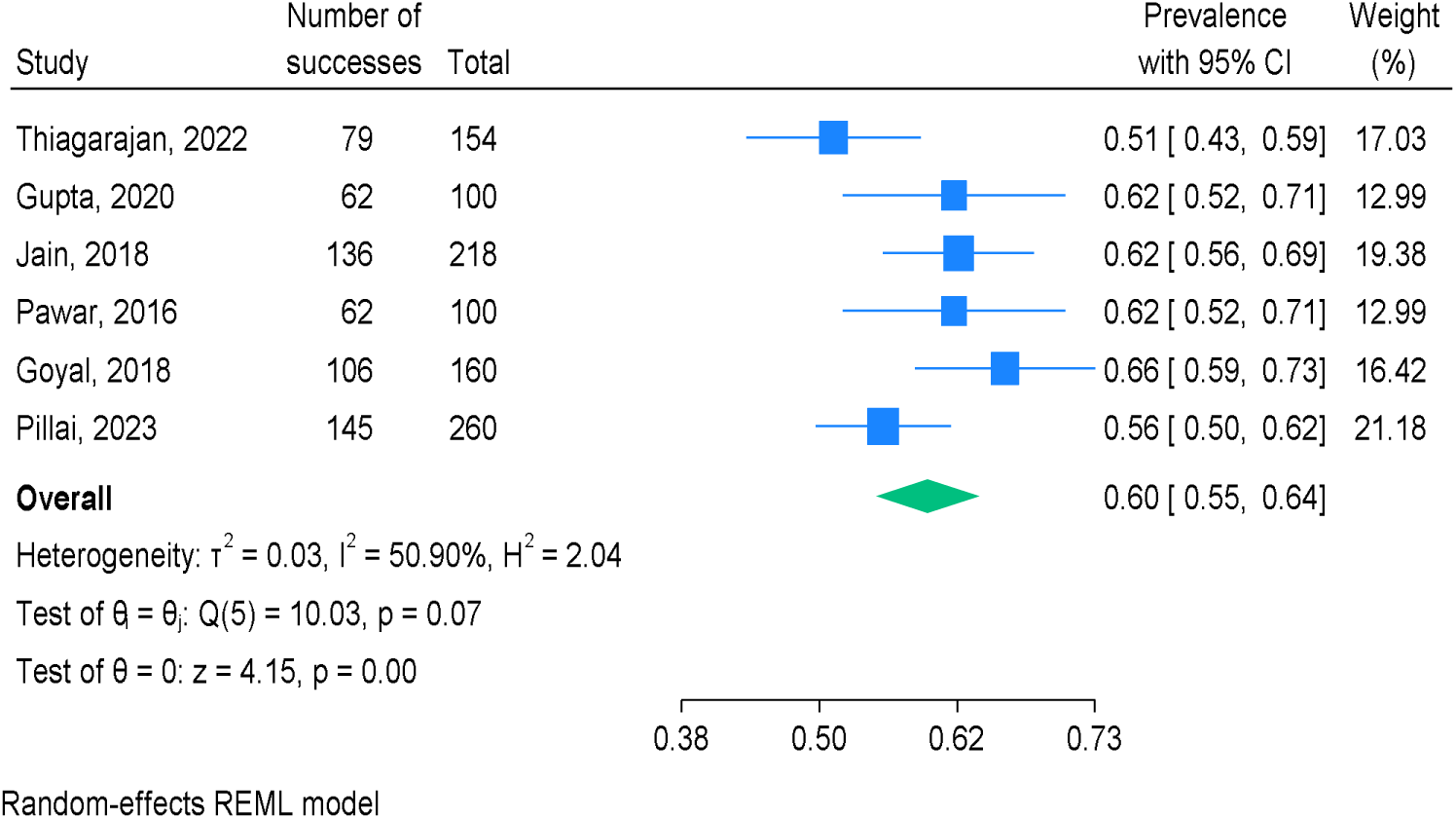
Pooled prevalence estimates of NAFLD in Overweight/obese Children and Adolescents(n=6)

### Assessment of methodological quality/Risk of bias Assessment

Joanna Brigg’s Institute (JBI) critical appraisal tool was used to assess the quality of the included studies (Fig 6). All studies had adequate response rates and the analysis performed with sufficient coverage of the identified sample i.e. children (n=11). Major issue in the quality of the included studies was inappropriateness of the sample frame(n=6) and lack of proper sampling technique(n=7).

**Fig. 6.**
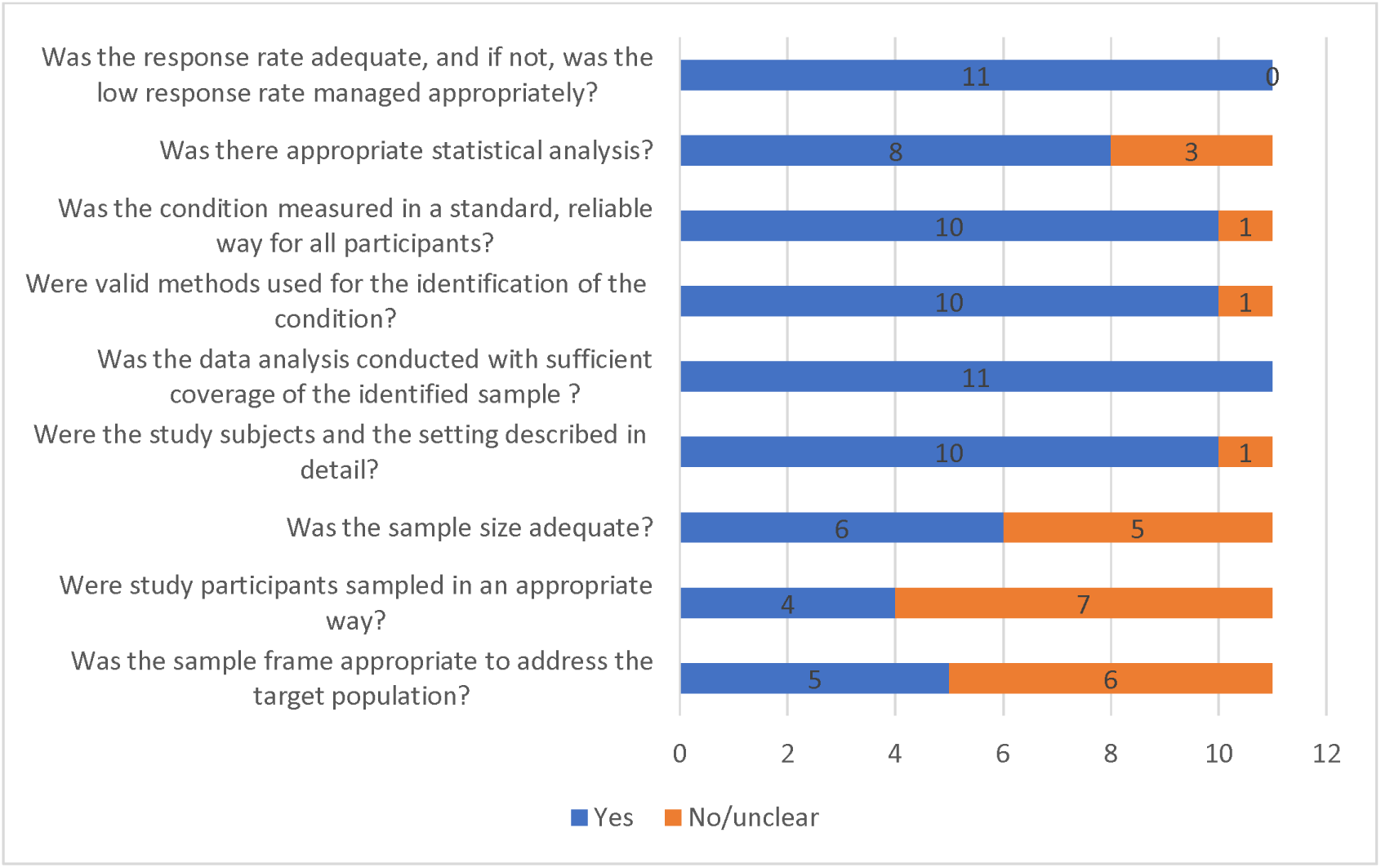
Assessment of study quality/risk of bias among the included studies using the JBI checklist.

**Fig. 7.**
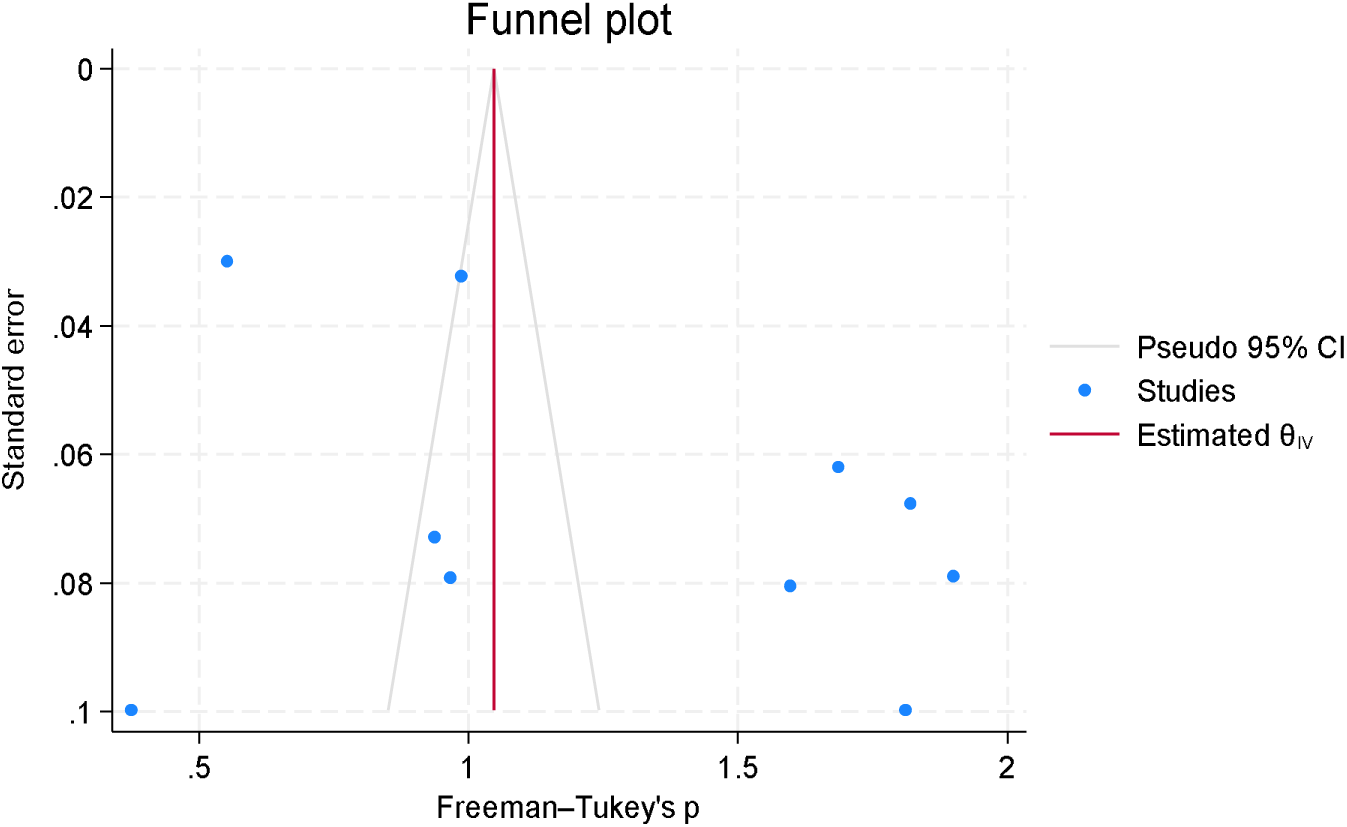
Funnel plot of the included studies Discussion.

Assessment using New Castle Ottawa Checklist ranged from four (Mehreen et al. (28)) to seven stars (Goyal et al. (23)) to the included studies out of maximum of seven stars with average star rating of 5.7 indicating good quality of the studies (Table 2).

**Table 2.**
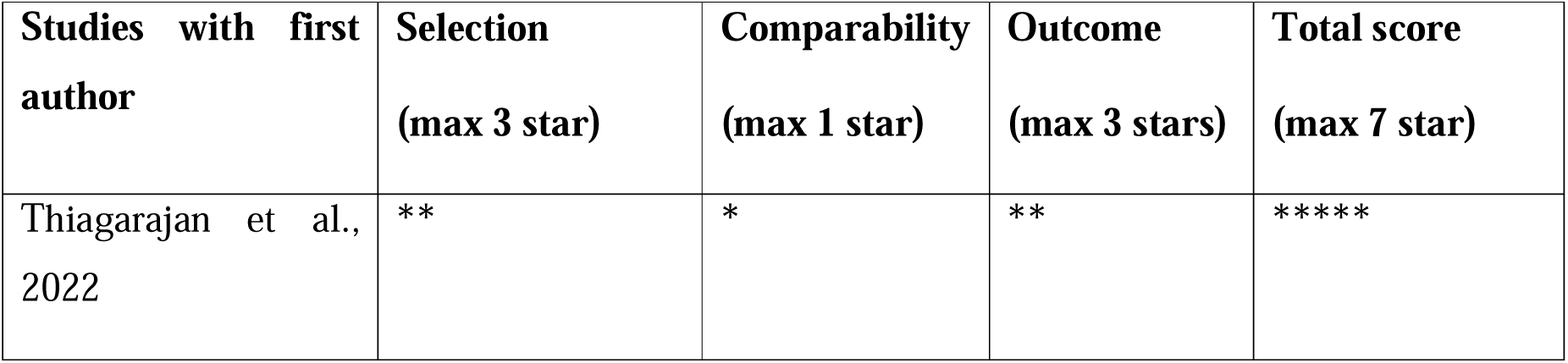
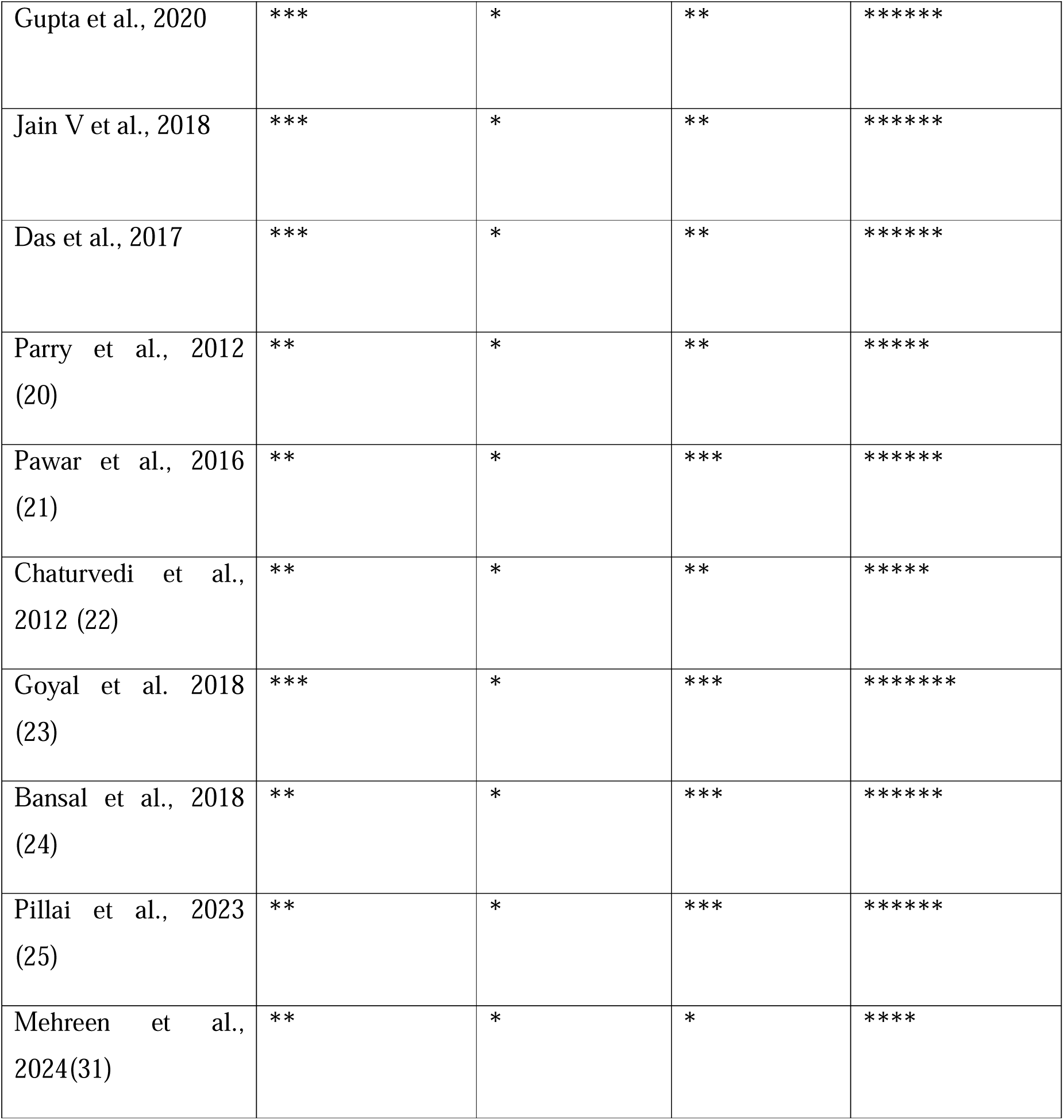
Quality assessment of the studies using New Castle Ottawa Scale for cross sectional studies.

### Publication Bias

Visual analysis of the funnel plot plotted for the included studies didn’t show marked asymmetry, implying non-significant publication bias. Similarly, regression-based Egger’s test for small-study effects using the random-effects model didn’t show a small-study effect (*p* = 0.32).

By pooling data from 11 studies with a combined total of 3512 participants, our meta-analysis estimated an overall NAFLD prevalence of 35.4% (95% CI: 19.7% -52.9 %). Subgroup analysis revealed that the prevalence among children from general populations was 10.7% (95% CI: 5.2%–20.5%), compared to a significantly higher prevalence of 59.7% (95% CI: 55.2%–64.1%) in groups identified as high-risk based on factors such as overweight and obesity. These findings have important implications for clinical practice and public health policy in India.

### Interpretation and Comparison with Previous Research

The high overall prevalence of NAFLD observed in our review is a cause of major public health concern, particularly as it reflects the increasing burden of paediatric obesity and related metabolic disorders in India. Similar to the findings of Neri et al., who highlighted liver steatosis as a significant metabolic risk marker in children, our results demonstrating a strong correlation between excess body weight and NAFLD among Indian children and adolescents (32). This association is further supported by Raj et al., who reported that adolescent obesity, sedentary lifestyle, and poor dietary patterns are key contributors to NAFLD development. This association is particularly concerning in the context of India, where rapid urbanization, reduced physical activity, and significant shifts in dietary patterns have contributed to an unprecedented rise in childhood obesity (33). The present findings are in line with those of Goel et al. and Minnu et al., both of whom highlighted how rapid urbanization, decreased physical activity, and a shift toward high-calorie, processed foods have led to a rise in childhood obesity—a primary driver of NAFLD (34, 35). When we compare our findings against studies conducted in other geographic regions or within adult populations, it becomes evident that Indian children—especially those with identifiable risk factors such as overweight and obesity—are disproportionately affected by NAFLD. Our results imply that early metabolic alterations, potentially stemming from lifestyle changes, lead to hepatic lipid accumulation long before the onset of overt clinical manifestations. Our data reflect these trends, with a substantially higher prevalence of NAFLD among high-risk groups such as overweight and obese children. Brizawasi et al. also emphasized that early metabolic disruptions can predispose children to fatty liver disease even before overt clinical symptoms arise, a pattern consistent with our pooled results. When compared to studies conducted in adult populations, such as the work by Shalimar et al., who found a high burden of NAFLD among Indian adults, our findings suggest that the disease burden begins much earlier, particularly in individuals with identifiable risk factors. Similar to global findings by Wong et al., who observed a shift in NAFLD epidemiology affecting younger populations, our results show that Indian children are not exempt from this trend and may, in fact, be disproportionately affected due to compounding risk factors. While Pawar et al. demonstrated a high prevalence of NAFLD (over 60%) among overweight Indian children, our review strengthens this evidence by aggregating data from multiple studies across regions, confirming that this is not an isolated observation but a national concern (15, 24, 36, 37). This early burden not only increases the risk of progression to non-alcoholic steatohepatitis (NASH) but also raises concerns about the long-term implications for cardiovascular health and overall mortality (38).

It is important to note that while there was a consistent trend showing higher NAFLD prevalence in high-risk groups across the included studies, considerable heterogeneity was observed (with I² values approaching 99% overall). This variability likely reflects several critical factors: differences in study design, regional variations in population characteristics, and inconsistencies in the diagnostic criteria employed (12). For instance, while most studies relied on ultrasonography as a primary tool for diagnosis, the sensitivity and specificity of this method can vary markedly between settings and operators. These diagnostic discrepancies, combined with variations in sample selection and study protocols, contribute to the overall heterogeneity of the results (39).

Despite these variations, the repeated observation of elevated NAFLD prevalence in high-risk paediatric populations aligns with global research that establishes a clear connection between obesity and hepatic fat accumulation. This consistency across studies underscores the urgency for implementing early screening programs (40, 41). By identifying at-risk children early on, healthcare providers have an opportunity to intervene with tailored lifestyle and dietary modifications before more severe hepatic pathology develops(42, 43). Ultimately, such proactive measures could play a crucial role in curbing the progression of NAFLD and its related complications, thereby reducing the future burden on the healthcare system.

In summary, our interpretation of the findings not only brings attention to the urgent challenge of NAFLD among Indian children, particularly those who are overweight or obese, but also reinforces the need for standardized diagnostic protocols and timely intervention strategies. The insights from our review contribute to the broader discourse on paediatric NAFLD and support the call for comprehensive, early-stage preventive measures within this vulnerable population.

### Clinical and Public Health Implications

The results of this systematic review carry significant implications for both clinical practice and public health strategies. The elevated prevalence of NAFLD among Indian children and adolescents, especially within high-risk populations such as those with obesity, points to a growing yet under-recognized burden of liver disease in the paediatric population. This highlights the need for the integration of NAFLD screening and management into standard paediatric care protocols (44, 45).

From a clinical perspective, the findings suggest that routine liver health assessments should be considered for children who present with risk factors such as overweight, obesity, or features of metabolic syndrome. These assessments could include non-invasive imaging modalities like ultrasonography, alongside liver function tests, as part of routine check-ups in paediatric clinics. Early identification of hepatic steatosis offers an opportunity for timely intervention, which is critical given the potential progression of NAFLD to more advanced liver diseases such as non-alcoholic steatohepatitis (NASH), fibrosis, cirrhosis, and even hepatocellular carcinoma if left untreated (45, 46).

Additionally, clinicians should be encouraged to adopt a multidisciplinary approach to managing NAFLD in children. This includes not only hepatological evaluation but also nutritional counselling, physical activity recommendations, and psychological support when necessary. Empowering families with knowledge about lifestyle changes—such as adopting a balanced diet, reducing screen time, and increasing physical activity—can be instrumental in managing and even reversing early-stage NAFLD in paediatric patients (47, 48).

From a public health perspective, the findings advocate for a shift toward preventive, community-focused strategies aimed at addressing the root causes of paediatric NAFLD. Community-based screening programs, particularly in schools or primary health centres, can serve as effective tools for identifying at-risk children early. These programs can be integrated into existing child health initiatives or school health programs, ensuring broader coverage and sustainability (44).

Furthermore, the growing trend of childhood obesity and sedentary behaviour in India calls for the development and implementation of national and regional policies that promote healthy growth and metabolic well-being in children. Public health campaigns focusing on nutrition education, physical activity promotion, and parental awareness can play a critical role in shifting societal behaviors. Regulatory measures, such as limiting the advertisement of unhealthy foods to children or improving the nutritional quality of school meals, may also support long-term prevention efforts (42).

In addition, collaboration between stakeholders—including healthcare providers, educators, policymakers, and families—is vital to ensure that efforts to address paediatric NAFLD are comprehensive and sustained. By aligning clinical practices with broader public health initiatives, it is possible to create an ecosystem that supports early detection, risk reduction, and better long-term outcomes for children at risk of NAFLD (37, 45).

Screening programs conducted at the community level—especially within schools and primary health centres—can be valuable in the early identification of children at risk for NAFLD (41). By incorporating these initiatives into existing child health or school-based programs, it is possible to achieve wider reach and ensure long-term sustainability of such preventive efforts.

### Limitations

Despite the valuable insights provided by our meta-analysis, several limitations merit consideration. First, all 11 studies included were single-center investigations with relatively small sample sizes, which may limit the generalizability of our findings. The considerable heterogeneity observed across studies likely reflects variations in sample frames, diagnostic criteria, and study designs. Additionally, the predominant reliance on ultrasonography, which may have reduced sensitivity for mild steatosis, could have led to under- or overestimation of the true prevalence in some settings. Finally, potential biases related to study selection, regional variations, and sampling techniques are inherent in the reviewed literature, underscoring the need for more rigorous multicentric studies.

## Conclusion

In conclusion, this systematic review demonstrates that non-alcoholic fatty liver disease (NAFLD) poses a growing health concern among Indian children and adolescents, with a disproportionately high burden observed in those who are overweight or obese. The consistently elevated prevalence in high-risk groups underscores the urgent need for integrating early screening measures into routine paediatric healthcare, alongside implementing targeted lifestyle interventions. These findings also call for the development of comprehensive public health strategies focused on obesity prevention and metabolic health promotion from a young age. Addressing existing methodological limitations through well-designed, standardized, and longitudinal research will be essential in shaping effective clinical guidelines and public health policies. Such efforts are critical not only to prevent disease progression in affected individuals but also to reduce the long-term health and economic consequences associated with paediatric NAFLD in India.

## Supporting information

Supplemental Table 1 key words search result

## Data Availability

All data produced in the present work are contained in the manuscript

## Acknowledgements

Authors would like to acknowledge all the authors whose articles were included for this review

