## Supplemental Table 1 key words search result for "Prevalence of Non-alcoholic Fatty Liver Disease (NAFLD) among Children and Adolescents (<18 years) in India: A Systematic Review and Meta-analysis"

| **Database** | **Key terms** | **Number of articles retrieved** |
| --- | --- | --- |
| **PubMed** | ("non-alcoholic fatty liver disease"[All Fields] OR "nafld"[All Fields] OR "fatty liver disease"[All Fields] OR "nash"[All Fields] OR "fatty liver"[All Fields] OR ("fatty"[All Fields] AND "liver"[All Fields]) OR "steatohepatitis"[All Fields] OR ("non-alcoholic"[All Fields] AND "fatty"[All Fields] AND "liver"[All Fields] AND "disease"[All Fields])) AND ("child"[All Fields] OR "children"[All Fields] OR "paediatric"[All Fields] OR "pediatric"[All Fields] OR ("infant"[MeSH Terms] OR "infant"[All Fields] OR "infants"[All Fields] OR "infant s"[All Fields]) OR "adolescen*"[All Fields]) AND ("india"[MeSH Terms] OR "india"[All Fields] OR "india s"[All Fields] OR "indias"[All Fields] OR ("andaman"[All Fields] OR "andamans"[All Fields]) OR "Nicobar"[All Fields] OR "Andhra Pradesh"[All Fields] OR "Arunachal Pradesh"[All Fields] OR "Assam"[All Fields] OR "Bihar"[All Fields] OR "Chandigarh"[All Fields] OR "Chhattisgarh"[All Fields] OR "Dadar"[All Fields] OR "Daman"[All Fields] OR ("delhi"[All Fields] OR "delhi s"[All Fields]) OR "New Delhi"[All Fields] OR "Diu"[All Fields] OR "Goa"[All Fields] OR "Gujarat"[All Fields] OR "Haryana"[All Fields] OR "Haveli"[All Fields] OR "Nagar Haveli"[All Fields] OR "Himachal Pradesh"[All Fields] OR "Jammu"[All Fields] OR "Jharkhand"[All Fields] OR "Karnataka"[All Fields] OR "Kashmir"[All Fields] OR "Kerala"[All Fields] OR "Lakshadweep"[All Fields] OR "Madhya Pradesh"[All Fields] OR "Maharashtra"[All Fields] OR "Mumbai"[All Fields] OR "Bombay"[All Fields] OR "Manipur"[All Fields] OR "Meghalaya"[All Fields] OR "Mizoram"[All Fields] OR "Nagaland"[All Fields] OR "Nicobar"[All Fields] OR "Odisha"[All Fields] OR "Puducherry"[All Fields] OR "Pondicherry"[All Fields] OR "Punjab"[All Fields] OR "Rajasthan"[All Fields] OR ("sikkim"[MeSH Terms] OR "sikkim"[All Fields]) OR "Tamil Nadu"[All Fields] OR "Chennai"[All Fields] OR "Madras"[All Fields] OR "Telangana"[All Fields] OR "Tripura"[All Fields] OR "Uttar Pradesh"[All Fields] OR "Uttarakhand"[All Fields] OR "Uttaranchal"[All Fields] OR "West Bengal"[All Fields] OR ("bengal"[All Fields] OR "bengale"[All Fields] OR "bengals"[All Fields]) OR ("calcutta"[All Fields] OR "calcutta s"[All Fields])) | 222 |
| **Scopus** | (TITLE-ABS-KEY ( "non-alcoholic fatty liver disease" OR "fatty liver disease" OR nash OR steatohepatitis OR "fatty liver" OR nafld ) AND TITLE-ABS-KEY ( "child" OR "children" OR "paediatric" OR "pediatric" OR infant OR "adolescen*" ) AND TITLE-ABS-KEY ( india OR andaman OR nicobar OR "Andhra Pradesh" OR "Arunachal Pradesh" OR assam OR bihar OR chandigarh OR chhattisgarh OR dadar OR daman OR delhi OR "New Delhi" OR diu OR goa OR gujarat OR haryana OR haveli OR "Nagar Haveli" ) ) | 145 |
| **Embase** | "non-alcoholic fatty liver disease" OR "nafld" OR"fatty liver disease" OR "nash" OR "fatty liver" OR ("fatty" AND "liver") OR "steatohepatitis" OR ("non alcoholic"AND "fatty" AND "liver" AND "disease") **AND** ("child" OR “children" OR "paediatric" OR "pediatric" OR infant OR "adolescen*") **AND** ((India OR Andaman OR Nicobar OR "Andhra Pradesh" OR "Arunachal Pradesh" OR Assam OR Bihar OR Chandigarh OR Chhattisgarh OR Dadar OR Daman OR Delhi OR "New Delhi" OR Diu OR Goa OR Gujarat OR Haryana OR Haveli OR "Nagar Haveli" OR "Himachal Pradesh" OR Jammu OR Jharkhand OR Karnataka OR Kashmir OR Kerala OR Lakshadweep OR "Madhya Pradesh" OR Maharashtra OR Mumbai OR Bombay OR Manipur OR Meghalaya OR Mizoram OR Nagaland OR Nicobar OR Odisha OR Puducherry OR Pondicherry OR Punjab OR Rajasthan OR Sikkim OR "Tamil Nadu" OR Chennai OR Madras OR Telangana OR Tripura OR "Uttar Pradesh" OR Uttarakhand OR Uttaranchal OR "West Bengal" OR Bengal OR Calcutta)) | 125 |
| **CINAHL** | **PubMed/Medline**- "non-alcoholic fatty liver disease" OR "nafld" OR"fatty liver disease" OR "nash" OR "fatty liver" OR ("fatty" AND "liver") OR "steatohepatitis" OR ("non-alcoholic” AND "fatty" AND "liver" AND "disease") **AND** ("child" OR “children" OR "paediatric" OR "pediatric" OR infant OR "adolescen*") **AND** ((India OR Andaman OR Nicobar OR "Andhra Pradesh" OR "Arunachal Pradesh" OR Assam OR Bihar OR Chandigarh OR Chhattisgarh OR Dadar OR Daman OR Delhi OR "New Delhi" OR Diu OR Goa OR Gujarat OR Haryana OR Haveli OR "Nagar Haveli" OR "Himachal Pradesh" OR Jammu OR Jharkhand OR Karnataka OR Kashmir OR Kerala OR Lakshadweep OR "Madhya Pradesh" OR Maharashtra OR Mumbai OR Bombay OR Manipur OR Meghalaya OR Mizoram OR Nagaland OR Nicobar OR Odisha OR Puducherry OR Pondicherry OR Punjab OR Rajasthan OR Sikkim OR "Tamil Nadu" OR Chennai OR Madras OR Telangana OR Tripura OR "Uttar Pradesh" OR Uttarakhand OR Uttaranchal OR "West Bengal" OR Bengal OR Calcutta)) | 315 |
